# Impact assessment of accreditation in primary and secondary public Healthcare Institutions in the State of Kerala, India

**DOI:** 10.1101/2020.05.30.20117432

**Authors:** Sindhu Joseph

## Abstract

**Objectives:** This study examines the impact of accreditation on the quality of public healthcare delivery in primary and secondary healthcare facilities in Kerala, India.

**Study Design:** A cross-sectional study.

**Setting:** Kerala State, India

**Participants:** Participants are the in-patients (621) who are admitted in medical wards at accredited (312) and non-accredited (309) public healthcare facilities.

**Main Outcome Measures:** Ten constructs used in the study, overarching the quality healthcare delivery, adapting previous studies, SERVQUAL, and Donabedian’s SPO models, are Physical Facility, Admission Services, Patient centeredness, Accessibility of Medical Care, Financial Factors, Professionalism, Staff Services, Medical Quality, Diagnostic Services, and Patient Satisfaction.

**Methods:** The study employed a positivist approach using a survey questionnaire. The study was conducted from July 2017 to July 2018, using stratified random sampling consists of the four strata; GHs, W&C hospitals, THQHs/THs and CHCs.

**Results:** Accreditation has a positive impact on patient satisfaction and other quality dimensions, overarching structural and procedural quality in primary healthcare facilities under the public sector in Kerala. Conversely, accreditation has not improved the quality dimensions in secondary healthcare facilities and thereby, the satisfaction of patients.

**Conclusions:** It cannot be assumed that accreditation is always associated with quality care in primary healthcare facilities. The implementation process must be systematic and must be regularly monitored to make it useful. Mere structural transformation through accreditation alone cannot guarantee patient satisfaction. Secondary healthcare facilities must be transformed into quality care centres through rhetoric action of the authorities concerned through organized efforts.

## 1. INTRODUCTION

The Healthcare system has a mirror image on the socio-economic development of society. Quality of healthcare has been a widely discussed domain, and many institutions and organizations perceive accreditation as a useful tool for quality transformation. Accreditation had its early beginnings in the USA, where Earnest Codman introduced the ‘end result system’ in 1910, insisting hospitals to track each patient to test the effectiveness of healthcare delivery(1). Globally, since the 1970s, healthcare quality improvement activities are progressing through healthcare accreditation programs and accrediting organizations(2,3). Accreditation is a means of publicly recognizing a healthcare organization against predetermined performance standards of operation by trained external peer reviewers(4–8) and encourages the development of professional skills, cost management, increased structure, efficient management of care, and appreciation among workers(9). Accreditation enables one to have an introspection based on the reports and recommendations of the accreditation team and, therefore, enables them to benchmark themselves(10).

Accreditation benefits all healthcare stakeholders, such as medical and paramedical professionals, patients, and the public as a whole(10). Standardization of cost and quality is not enforced in the present healthcare system, and therefore, accreditation is the only probable means of proclaiming healthcare reliability and authenticity. It provides public recognition and develops trust between users and service providers(6,8,11). To summarize, accreditation can be perceived as a measure to maintain and improve quality, public safety, legal recognition, risk management, private sector monitoring, implementation of new delivery settings, address national public health issues, the formation of new systems or networks of services and create centers of excellence(7).

Accreditation programs are implemented at international (Joint Commission International-JCI), national (for instance, National Accreditation Board for Hospitals & Healthcare Providers (NABH-India) and state levels (for example, Kerala (Kerala Accreditation Standards for Hospitals-KASH).

### 1.1 Accreditation and Indian Healthcare

Accreditation has become an essential parameter for the private healthcare sector in India, and hence, accreditation programs have been implemented voluntarily, especially to cater to medical tourists, and this progressive transformation is visible now in the public sector too. Indian accreditation drive was initiated in the 1930s, including nursing homes and hospitals under one umbrella(1). To accredit hospitals at the national level, NABH is set up with full functional autonomy in its operation and working at par with global benchmarks and is being supported by all stakeholders, including industry, consumers, government. Kerala has the largest number of NABH accredited hospitals in India under the public sector(12).

#### 1.1.1 Healthcare of Kerala

Kerala healthcare model is accepted globally, especially after the successful prevention and treatment against the epidemic Nipah and the pandemic COVID-19. The healthcare status of Kerala is on par with the western countries(12). In 2011, Kerala achieved the highest Human Development Index (HDI) in India, based on its performance in key measures (Mukherjee and Chakraborty, 2014). Kerala’s ‘Infant Mortality Rate (IMR) and ‘Maternal Mortality’ Rate (MMR)’ (Table 1) are comparable with global standards(13). These quality indicators are the cumulative result of ‘both supply-side interventions’ by political administrations and other agencies, and ‘demand-side interventions by social movements’(14). It is found that in 2017–18, public sector medical facilities are availed by 48 percent of the population, which was 34 percent in 2014 (NSSO 2015; NSO 2019 in(14)).

#### 1.1.2 Primary and Secondary Healthcare in Kerala

Kerala public healthcare system functions decentralized under a three-tier concept envisaged delivering primarily to specialty healthcare. At the tertiary level, Medical Colleges provide specialty services on a ‘referral basis’ whereas District Hospitals (DHs), General Hospitals (GHs), Women & Children (W&C) hospitals and the Taluk Head Quarters /Taluk Hospitals (THQHs/THs provide secondary care(15). The Community Health Centers (CHCs) at the block level represent the primary care hospitals along with Primary Health Centers (PHCs) at Grama Panchayat (village) level which operate as ambulatory care providers, health education, immunization, family planning services, counseling, health awareness education, social security, and rehabilitation(15) to achieve the goal ‘Health for all’ (Park, 2009). Currently, 168 PHCs are designated as “family health centers that function round the clock (PHCs 24×7) and provide free treatment for lifestyle diseases”(16).

Kerala public healthcare infrastructure includes 848 PHCs, 232 CHCs, 81 THQHs/ THs, 18 DHs, and 18GHs (as on 31.03.2017) ((17), p.8). For the quality enhancement, the 13th Five-Year Plan of Kerala focused on the implementation of the specialty cadre in all health care institutions up to the level of CHCs and modernization of the functioning of the PHCs as Family Health Centers(12). One PHC serves for every 30000 population (20000 population in Tribal and Hilly area), one CHC for every 120000 population (80000 population in Tribal and Hilly area), THQH/THS for 200000 population and one GH/DH for entire district population(18).

Kerala’s primary care system became a model as seven PHCs were awarded the National Quality Assurance Standards (NQAS) certification from National Health Mission, Ministry of Health and Family Welfare (India), in 2017(19). The status of ‘top performer and best PHC’ in the country was bagged by Kayyur PHC in Kasaragod district in Kerala, on the basis of its infrastructural services, maintenance, and patient-friendliness(19). Despite the domination of the private sector in the Kerala healthcare industry, a large number of its 33.3 million population, especially the poor with a monthly income of less than Rs. 5000 and a group of vulnerable populations of women and children seek healthcare from public sector hospitals(20).

Primary care has been dramatically improved by the State involving the local governments which take the lead in infrastructure development of PHCs and sub-centers, purchase of medical equipment and drugs, filling up of vacancies on contract, and also supplementing the honorarium of ASHA (Accredited Social Health Activist) workers(14).

#### 1.1.3 Accreditation and Kerala healthcare system

Currently, according to the NABH website, Kerala has 34 private hospitals and five public hospitals accredited by NABH and 3 JCI accredited hospitals when compared with 2005 when there was no single hospital accredited. Being a newly introduced state-level accreditation program in 2012, KASH aims to uplift the quality standards and services given by the government hospitals in all care settings with a modest investment. After the achievement of KASH, the individual hospitals may opt for higher standards viz. NABH, which requires more investment and effort(12,21).

### 1.2 Impact of accreditation on quality enhancement and patient satisfaction

It is important to advance the knowledge on accreditation impact in hospital settings, especially on the structural, procedural, and outcome levels, when considering the financial investment, effort, and time to attain quality assurance certifications. Assessment of impact is important since it has become a common platform of quality transformation for primary, secondary, and tertiary care systems(6). Many conceptual and empirical studies focused on the theme of accreditation in varied settings and space. Accreditation and quality enhancements(4,22,23), and thereby, patient satisfaction is correlated positively by many researchers(9,24–30). Despotou et al.,(31), Carlson-Stevens et al.,(32) and Turrioni et al.,(22) confirmed the positive impact of accreditation in tertiary care in South Korea, India, and Brazil respectively. Ghareeb, Said & El Zoghbi(33) found a positive impact of accreditation in the primary healthcare settings in Qatar. Al Tehewy(24) and Williams et al. (34) demonstrated the positive effect accreditation on patient satisfaction and performance levels in their comparative studies at accredited and non-accredited settings.

Nonetheless, many other studies gave contradictory results on the accreditation impact(35–42), and few studies focused on the perception of personnel at different capacities in accredited hospitals(for instance, nurses) and on various accredited facilities (for example, ambulatory services) (32). Otaibi et al. (43) found the reverse effect of CBAHI (Saudi Central Board for Accreditation of Healthcare Institutions) on patient safety and failed to create total quality management.

Considering the previous studies and their inconsistent results, there is a need to assess the overall effectiveness of accreditation on quality. Further, there is a dearth of studies assessing the impact of accreditation while having a comparison group and from the patients’ perspective. Most importantly, the impact of accreditation programs in secondary healthcare care settings lacks evidence in the literature. In this backdrop, this study aimed to fill these gaps by assessing the impacts of accreditation implementation in primary and secondary public healthcare settings of Kerala while having a comparison group, from the patients’ perspective. The study results will have implications at the policy level and service provider level to restructure the implementation process of accreditation. To this end, this study sets the following objectives:

1. To measure the level of patient satisfaction in accredited and non-accredited primary and secondary public healthcare facilities in Kerala
2. To analyze the structural and procedural dimensions of healthcare delivery in accredited and non-accredited primary and secondary public healthcare facilities in Kerala

## 2. METHODS

A cross-sectional study drawn on a positivist approach was conducted from July 2017 to July 2018 using a questionnaire. The research used stratified random sampling where four strata, GHs, W&C hospitals, THs/THQHs (Secondary Healthcare Facilities) and CHCs (Primary Healthcare Facilities), are selected randomly from Trivandrum region representing Southern Kerala, Ernakulam region representing Central Kerala and Kozhikode region representing Northern Kerala to collect samples. The study’s target population was in-patients admitted to medical wards at accredited and non-accredited public hospitals in Kerala. Being primary care facilities, PHCs were excluded from the study because of the lack of an adequate number of in-patients available.

To get an authentic number of samples, 10% of the number of beds from each stratum was included in the study except GHs (15%), where there was only one GH accredited in Kerala (Table 2). A total of 760 questionnaires were distributed in the Inpatient wards, of which 621 (82%) were valid for analysis (312 from accredited and 309 from non-accredited), which is considered sufficient to represent a large population(44). In-patients aged 16 years or older and able to speak Malayalam or the English language were included in the study.

**Table 1:**

| Accredited Hospitals (NABH + CASH) |  |  |  | Non-accredited Hospitals |  |  |  |
| --- | --- | --- | --- | --- | --- | --- | --- |
| Hospital Type | Total no. of beds | Strata size | Sample size | Hospital Type | Total no. of beds | Strata size | Sample size |
| CHC | 6567 | 10% | 65 | CHC | 6567 | 10% | 65 |
| GH | 6920 | 15% | 103 | GH | 6920 | 15% | 106 |
| THQH/<br>TH | 8553 | 10% | 85 | THQH/<br>TH | 8553 | 10% | 85 |
| W & C | 5662 | 10% | 59 | W & C | 5662 | 10% | 53 |
| Total |  |  | 312 | Total |  |  | 309 |
| Total: 621 |  |  |  |  |  |  |  |

**Table 2:**

| **Sl. No.** | **Health Indicators** | **Kerala** | **India** |
| --- | --- | --- | --- |
| 1 | Birth rate ('000 population) | 14.8 | 22.1 |
| 2 | Death rate ('0000 population) | 7 | 7.2 |
| 3 | Infant mortality rate ('0000 population) | 13 | 47 |
| 4 | Child mortality rate 0-4 years ('0000 population) | 2 | 15 |
| 5 | Maternal mortality rate (per 100,000 live births) | 81 | 212 |
| 6 | Total fertility rate (children per woman) | 1.7 | 2.6 |
| 7 | Couple protection rate (in percent) | 62.3 | 52 |
| 8 | Life at birth (male) | 71.4 | 62.6 |
| 9 | Life at birth (female) | 76.3 | 64.2 |
| **Source: Directorate of Health Services** |  |  |  |

The questionnaire included 60 items in two sections. The first section sought demographic information on age, gender, educational level, marital status, employment status, and the reason for hospital selection. The second section measured patient’s views on healthcare received by them on a 5-point scale (1 = Strongly Agree to 5 = Strongly Disagree) using ten constructs adapting previous critical studies and models in the area(45–50). The five dimensions of SERVQUAL model (Tangibility, Reliability, Responsiveness, Assurance, and Empathy) and SPO (Structure-Process-Outcome) model of Donabedian are imbibed in the chosen constructs. The Donabedian model includes *structure* (the context in which care is delivered) including infrastructure, medicine availability, staff, financial factors and equipment; *Process* (denotes the transactions between patients and healthcare providers) including patient-centeredness and relationship dimensions; and *Outcomes* (denotes the effects of healthcare) including patient experience and satisfaction(51–53). The variables were grouped under ten constructs, namely Physical Facility (14 items), Admission Services (2 items), Patient centeredness (7 items), Accessibility of Medical Care (5 items), Financial Factors (5 items), Professionalism (4 items), Staff Services (4 items), Medical Quality (4 items), Diagnostic Services (2 items) and Patient Satisfaction (5 items).

The questionnaire was initially developed in the English language and subsequently translated into Malayalam. Based on the result from the pilot test, few questions were omitted from the questionnaire, in which respondents generally did not respond. The validity of the questionnaire was evaluated based on content validity and experts’ opinion. Cronbach’s Alpha value was higher than the guideline value of 0.6. Simple statistical techniques such as descriptive statistics, t-test, and Kruskal Wallis tests have been undertaken for analyzing data.

## 3. RESULTS

### 3.1 Dimensions of Healthcare Delivery

The measures to study the quality healthcare dimensions were Physical Facilities (PF), Admission Services (AS), Patient Centeredness (PC), Accessibility to Medical Care (AM), Financial Factors (FF), Professionalism (P), Staff Services (SS), Medical Quality (MQ) and Diagnostic Services (DS).

#### 3.1.1 Patient Satisfaction (*Outcome* Domain)

*The outcome* of healthcare delivery is measured by the Patient Satisfaction construct. As seen in Table. 3, accreditation impacted in-patient satisfaction in accredited primary care facilities (CHCs (*M* = 4.6 ± 0.41154) and THQH/TH category (*M* = 4.14 ± 0.90281) under the secondary healthcare facility. While in other secondary facilities (GH and W&C), the non-accredited category gets higher scores (W&C Hospitals (*M* = 4.43 ± 0.52094) and GH (*M* = 4.34 ± 1.2963). It shows that accreditation has a positive impact on patient satisfaction in primary healthcare facilities but not in secondary care facilities.

**Table 3:**

| Institution Type | Accreditation Type | Hospital Type | Mean | Std. Deviation | Variance |
| --- | --- | --- | --- | --- | --- |
| Primary | Accredited | CHC | 4.60 | 0.41154 | 0.169 |
|  | Non-Accredited | CHC | 4.25 | 0.47398 | 0.225 |
| Secondary | Accredited | THQH/<br>TH | 4.29 | 1.41201 | 1.994 |
|  |  | GH | 4.27 | 0.59497 | 0.354 |
|  |  | W&C | 3.95 | 0.78199 | 0.612 |
| Secondary | Non-accredited | THQH/<br>TH | 4.14 | 0.90281 | 0.815 |
|  |  | GH | 4.34 | 1.29637 | 1.681 |
|  |  | W&C | 4.43 | 0.52094 | 0.271 |
| Primary Care Facility - CHCs |  |  | Secondary Care Facility - GH, W&C |  |  |

#### 3.1.2 Dimensions of Healthcare Delivery (*structure* domain)

*Structure* domain includes Physical Facility, Financial Factors, Staff Services, and Diagnostic Services.

##### 3.1.2.1 Healthcare delivery – Primary care facilities

The structure domain includes Physical Facility, Financial Factors, Staff Services, and Diagnostic Services. Table.4 shows that the accredited CHCs have been improved considerably in *structure* domain after accreditation (PF, M = 4.26; FF, M = 3.68; SS, M = 4.48; and DS, M = 4.23) when compared to non-accredited facilities (PF, M = 3.49; FF, M = 3.40; SS, M = 4.14; and DS, M = 2.78)

**Table 4:** Healthcare delivery dimensions at primary care hospitals-*Structure* Domain

| **Hospitals - Type** | **Stat** | **PF.** | **FF** | **SS** | **DS** |
| --- | --- | --- | --- | --- | --- |
| N-CHC | Mean | 3.49 | 3.40 | 4.14 | 2.78 |
|  | SD | 0.73 | 0.66 | 0.62 | 1.16 |
| A-CHC | Mean | 4.26 | 3.68 | 4.48 | 4.23 |
|  | SD | 0.49 | 0.69 | 0.60 | 0.72 |

##### 3.1.2.2 Healthcare delivery – secondary care facilities

As seen in Table.5, accreditation has impacted in THQH/TH with a higher score for all the four constructs (PF, M = 4.45; FF, M = 3.92; SS, M = 4.55; and DS, M = 4.43). Nonetheless, accredited GH and W&C hospitals receive a lesser score than the accredited. It shows that accreditation has impacted only at the lowest level in the secondary care facilities, in the structure domain, but not in the district level and specialty level hospitals.

**Table 5.**
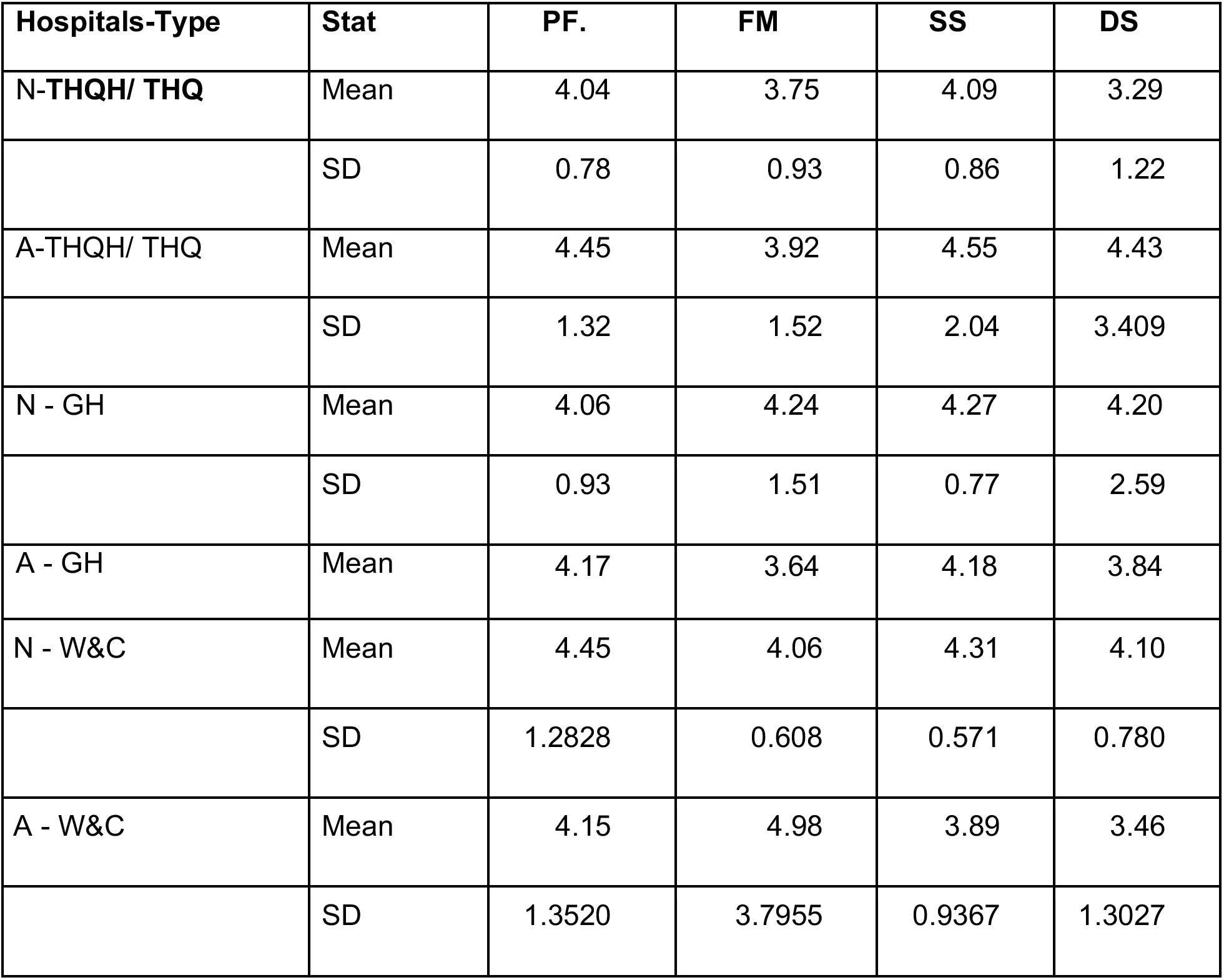
Healthcare delivery dimensions at Secondary care hospitals-*structure* Domain

#### 3.1.3 Dimensions of Healthcare Delivery *– Process* domain

The *process* domain includes Admission Services, Patient centeredness, Accessibility of Medical Care, Professionalism, and Medical Quality.

##### 3.1.3.1 Healthcare delivery – primary care facilities

Table.6 shows that the accredited CHCs have been improved considerably in the *process* domain (AS, M = 4.56; PC, M = 4.67; AM, M = 4.15; P, M = 4.36 and MQ, M = 4.53) when compared to non-accredited facilities (AS, M = 4.21; PC, M = 3.96; AM, M = 3.43; P, M = 3.95; and MQ, M = 4.40)

**Table 6:**

| Hospitals - Type | Stat | AS | PC | AM | P | MQ |
| --- | --- | --- | --- | --- | --- | --- |
| N-CHC | Mean | 4.21 | 3.96 | 3.43 | 3.95 | 4.40 |
|  | SD | 0.70 | 0.63 | 0.83 | 0.81 | 0.49 |
| A-CHC | Mean | 4.56 | 4.67 | 4.15 | 4.36 | 4.53 |
|  | SD | 0.54 | 0.35 | 0.97 | 0.88 | 0.51 |

##### 3.1.3.2 Healthcare delivery – secondary care facilities

As seen in Table.7, accreditation has impacted in THQH/TH marginally as two constructs out of four receive higher care in accredited facilities (AS, M = 4.72; PC, M = 4.26; AM, M = 3.32; P, M = 3.78; and MQ M = 4.61). In GH, both category hospitals receive almost identical scores. In W&C hospitals, all the four constructs except Admission Services receive a lesser score in accredited hospitals (AS, M = 4.53; PC, M = 4.15; AM, M = 3.67; P, M = 4.07; and MQ M = 4.35) than the non-accredited (AS, M = 4.07; PC, M = 4.43; AM, M = 4.12; P, M = 4.55; and MQ M = 4.74). Thus, it can be assumed that accreditation has not impacted the *process* domain.

**Table 7:** Healthcare delivery dimensions at secondary care hospitals-*process* domain

| Hospitals-Type | Stat | AS | PC | AM | P | MQ |
| --- | --- | --- | --- | --- | --- | --- |
| N-THQH/THQ | Mean | 4.72 | 4.26 | 3.32 | 3.78 | 4.61 |
|  | SD | 3.80 | 0.92 | 1.00 | 1.00 | 1.48 |
| A-THQH/THQ | Mean | 4.20 | 4.49 | 3.96 | 4.40 | 4.37 |
|  | SD | 0.85 | 1.01 | 1.27 | 2.12 | 0.701 |
| N-GH | Mean | 4.04 | 4.33 | 3.90 | 4.33 | 4.41 |
|  | SD | 0.89 | 0.67 | 0.99 | 1.16 | 0.74 |
| A-GH | Mean | 4.04 | 4.34 | 3.90 | 4.10 | 4.50 |
|  | SD | 0.94 | 1.00 | 0.92 | 0.81 | 1.435 |
| N-W&C | Mean | 4.07 | 4.43 | 4.12 | 4.55 | 4.74 |
|  | SD | 0.7990 | 0.522 | 0.5576 | 0.437 | 1.810 |
| A-W&C | Mean | 4.53 | 4.15 | 3.67 | 4.07 | 4.35 |
|  | SD | 0.9138 | 0.9137<br>4 | 1.2259 | 0.79607 | 0.515 |

## 4. DISCUSSION

This study was conducted to examine the impact of accreditation in primary and secondary healthcare facilities under the public sector in Kerala. The investigation constructs were subdivided into structural, procedural, and outcome dimensions according to the Donabedian model. The study implied that accreditation has brought a quality transformation in primary healthcare facilities in all these dimensions and hence confirms the Donabedian SPO model, which states that quality enhancement in structural, procedural aspects consequently results in patient satisfaction *(outcome)*. Further, this study confirms the findings of El Jardali et al. (54), who found that accreditation impacted the Lebanon primary care system. Similarly, Gharibi (33) noted that the USA, Australia, Canada, the UK, New Zealand, Jordan, Saudi Arabia, Lebanon, and Egypt (EMR) had well-developed and high-quality primary care accreditation models. The results of this study confirm the validity of global appreciation for the Kerala model for containing COVID-19 by effectively using its primary care centers with LSGs (Local Self Governments).

Despite the remarkable achievement in primary care facilities, the secondary care facilities fail to show any considerable changes in accreditation. Though the structure domain of THQH/TH receives a higher score in the accredited, all other facilities do not show any improvement in *structure* as well as the *process* domain. At the outcome level, THQHs/THs could trigger patient satisfaction because of accreditation, which is the next immediate referral point of CHCs. Nevertheless, patient satisfaction is less in accredited secondary care facilities, which are referral and specialist centers of healthcare. This outcome is to be considered seriously when satisfaction is ‘an expression of the patients’ overall judgment’(53) on ‘how well’ the services provided, and reflects patients’ perceptions’(36). Facilities intended to deliver more specialized services have not been impacted by accreditation, which contradicts the finding of Shaw(4), who found accreditation had benefitted larger hospitals in improving the quality of care. Reasons may be a limited supply of workforce, equipment and medicines, limited drug supplies and faulty equipment, high staff turnover, and workload, and the absence of a referral system makes the situation more challenging(54).

Despite the mixed response on the impact of accreditation in primary and secondary healthcare facilities, it is in line with the finding of El Jardali et al.(54), who noted that accreditation impacted hospitals with significant differences across hospital size. It can be assumed that accreditation is a step towards quality enhancement, especially in tangible terms(36); it is not necessarily a predictor of quality(40). This may be due to the misconception of the implementing agency that the patients will be content if the physical infrastructure is made appealing. The service characteristics of the healthcare aspect are often misconstrued, and consequently, the accreditation effort will be concentrated in improving tangible dimensions.

The results demonstrate four aspects. First, the success story of primary healthcare accreditation in the State and the other successful models (for instance, Jordanian, Egyptian, and Saudi Arabian) may be pursued before implementing accreditation at specialty and referral secondary care centers. Second, the Indian private healthcare sector has evolved mainly because of the incompetence of the government to provide basic services to its population(1). Therefore, financial botherations of the vulnerable population must be considered seriously as Maya(55) observed that average in-patient expense per person in the private hospital in Kerala is Rs.22, 989, whereas government sector cost is Rs.11,065 and the cost of an outpatient visit is Rs.525 in private and, Rs.391 in the public sector. Third, the leading health care transformation elements trust in ‘improved integration of care’ between the primary and secondary sectors(56). A ‘re-balancing of combined activity’ between primary and secondary (hospital) care, (57), requires a benchmarked indicator in quality dimensions. Kerala needs to equalize the two-tier systems in terms of quality and be integrated as another model for providing access to quality healthcare for all. Finally, Rahat (52) suggested that the hospitals must be empowered with more resources and knowledge “along with development and growth in determinants of quality in terms of structure, process, and outcome of the service’ before stepping into the accreditation process. If to receive the expected outcome from the accreditation process, there is a need for a rhetoric activity to transform the structural and procedural aspects of healthcare delivery.

Findings from this research may provide important insights for redefining priorities while implementing or planning to implement accreditation in Kerala or elsewhere. This study has implications for the entire healthcare industry, both public and private, and the policymakers in terms of providing visions in the strategic implementation of accreditation programs.

The data collection period witnessed the outbreak of Nipah fever in Kerala, and that affected the accessibility of data sources from the medical wards. Further, there was only one GH accredited hospital in Kerala, which might have affected the result. To honestly assess the impact of accreditation, the study requires more data from the same strata making the comparisons more valid, and hence the future research may include more data for the study to confirm these initial findings.

## 5. CONCLUSION

Accreditation can produce a positive repercussion in the hospital as a whole. Nonetheless, if to guarantee an expected outcome from this long and expensive process, the authorities must consider accreditation as a means of holistic and continuous transformation from conventional healthcare delivery. Hence, adequate human resource training must be imparted to deliver empathetic, patient-centered healthcare. Structure and process domains are the two sides of a coin and equally contribute to patient satisfaction. It has been suggested that the authorities have to intensify their monitoring and supervisory roles with an unwavering urge for excellence in the implementation process, to create an increased public acknowledgment.

## Data Availability

Data is confidential.

## Acknowledgement

The author thanks the Indian Council of Social Science Research (ICSSR), New Delhi for assisting this study through a Minor Research Project.

